# Contrasting epidemiology and population genetics of COVID-19 infections defined with 74 polymorphic loci in SARS-CoV-2 genomes sampled globally

**DOI:** 10.1101/2021.04.25.21255897

**Authors:** Felicia Chan, Ricardo Ataide, Jack S. Richards, Charles A. Narh

**Affiliations:** Central Clinical School, Monash University, Melbourne, Australia; Burnet Institute for Medical Research, VIC 3004, Melbourne, Australia; Department of Medicine, University of Melbourne, Australia; Walter and Eliza Hall Institute, Melbourne, Australia; Departmet of Infectious Diseases, Monash University, Melbourne, Australia

**Keywords:** COVID-19, SARS-CoV-2, epidemiology, genetics, multilocus-genotypes, evolution, linkage, mutation and transmission

## Abstract

SARS-CoV-2, the coronavirus causing COVID-19, has infected and killed several millions of people worldwide. Since the first COVID-19 outbreak in December 2019, SARS-CoV-2 has evolved with a few genetic variants associated with higher infectivity. We aimed to identify polymorphic loci in SARS-CoV-2 that can be used to define and monitor the viral epidemiology and population genetics in different geographical regions. Between December 2019 and September 2020, we sampled 5,959 SARS-CoV-2 genomes. More than 80% of the genomes sampled in Africa, Asia, Europe, North America, Oceania and South America were reportedly isolated from clinical infections in older patients, ≥ 20 years. We used the first indexed genome (NC_045512.2) as a reference and constructed multilocus genotypes (MLGs) for each sampled genome based on amino acids detected at 74 polymorphic loci located in ORF1ab, ORF3a, ORF8, matrix (M), nucleocapsid (N) and spike (S) genes. Eight of the 74 loci were informative in estimating the risk of carrying infections with mutant alleles among different age groups, gender and geographical regions. Four mutant alleles - ORF1ab L_4715_, S G_614_, and N K_203_ and R_204_ reached 90% prevalence globally, coinciding with peaks in transmission but not COVID-19 severity, from March to August 2020. During this period, the MLG genetic diversity was moderate in Asia, Oceania and North America; in contrast to Africa, Europe and South America, where lower genetic diversity and absence of linkage disequilibrium indicated clonal SARS-CoV-2 transmission. Despite close relatedness to Asian MLGs, MLGs in the global population were genetically differentiated by geographic region, suggesting structure in SARS-CoV-2 populations. Our findings demonstrate the utility of the 74 loci as a genetic tool to study and monitor SARS-CoV-2 transmission dynamics and evolution, which can inform future control interventions.

## Introduction

Knowledge of the epidemiology and transmission dynamics of SARS-CoV-2, the causative agent of the COVID-19 pandemic, is seminal to public health control efforts. SARS-CoV-2 has more than 110 million confirmed cases – which resulted in 2.5 million deaths – as of March 2021. Since the first outbreaks in China in December 2019, SARS-CoV-2 transmission hotspots have shifted spatio-temporally from Asia to Europe, followed by North America and South America [1, 2]. Testing and isolation of infected individuals have been integral, as public health interventions, to control the virus’ spread. Global reports have shown significant differences in the prevalence, distribution and demographics of COVID-19 cases; however, little is known about how these epidemiological differences relate to infection dynamics.

SARS-CoV-2 is a beta-coronavirus with a positive single-stranded RNA genome of ∼30 kb. It shares 80% of its genome with SARS-CoV, which caused the 2003-2004 SARS outbreak [3]. A prominent structural feature is the spike glycoprotein (S), which facilitates cell entry and is a target of host immune responses. Other structural proteins include the nucleocapsid (N), envelope (E) and matrix/membrane (M) proteins, which are involved in viral assembly and priming of host immune responses [4]. Nearly half of the genome comprises an opening reading frame 1ab (*orf1ab*), which encodes 16 non-structural proteins (NSP 1-16) that constitute the replicase machinery. Notable NSPs include NSP1 (suppresses host immune responses), NSP5 (encodes viral 3C-like protease) and NSP12 (encodes the RNA-dependent RNA polymerase, i.e., RdRP). Other ORFs, including ORF3a (induces apoptosis in host cells), ORF6, ORF7a, ORF8 (ORF8 mediates immune suppression and evasion) and ORF10 encode accessory proteins that are involved in viral replication and host immune dysregulation [4].

Since first being identified, the virus has evolved, with numerous genetic variants being associated with higher infectivity. The geographical distribution and probable risk factors (e.g., demographics and clinical factors) for infection with mutant genotypes remain unknown. Comparative genomic analysis of SARS-CoV-2 infections collected globally suggests that the virus is adapting to its human host. A few genetic variants harbouring E484K, N501Y and D614G mutations in the S protein have been associated with higher infectivity than the wild-type variant, Wuhan NC_045512.2 [5, 6]. Variants with these mutations rose to predominance in many parts of the United Kingdom and South Africa [7, 8]. Other mutations, including *orf1ab* P4715L, *Orf3a* G251V and *orf8* L84S, have been associated with higher infectivity and viral density, respectively [9]. Whether these and other unreported mutations are linked, under selection and can be used to source-track infections within and between different geographical regions has not been investigated.

More polymorphic and informative loci, representative of the global SARS-CoV-2 genetic diversity, are needed to accurately differentiate closely related variants and interrogate the virus population genetics in different geographical regions [9, 10]. Comparison of SARS-CoV-2 whole genomes identified phylogenetic clusters, defined by Single Nucleotide Polymorphisms (SNPs) in < 10 codons/loci, that differentiated European and Asian infections [11, 12]; however, these loci lacked the needed resolution to differentiate variants circulating globally. Multilocus genotyping using amino acid changes in SARS-CoV-2 can reduce the complexity in the genomic data and provide informative and virologically relevant data that can provide insights into the transmission dynamics and evolution of variants causing COVID-19. This approach on multiple polymorphic loci can estimate and monitor the genetic diversity of SARS-CoV-2 populations spatiotemporally and in response to control interventions.

This study evaluated the epidemiology and population genetics of 5,959 SARS-CoV-2 genomes sampled globally to identify risk factors associated with infection with mutant variants and gain insights into how the viral population had evolved geographically eight months into the pandemic. Briefly, we identified 74 polymorphic loci, of which eight loci located in *orf1ab, orf3a, orf8*, N and S genes, were considered informative in explaining the risk of infection with mutant variants among different demographics and COVID-19 disease phenotypes. Multilocus genotyping at the 74 loci allowed us to genetically differentiate closely related variants circulating globally and gain insights into the viral population genetics in different geographical regions.

## Material and Methods

### Data curation and study variables

The current study sought to investigate the epidemiology and population genetics of SARS-CoV-2 genetic variants causing the COVID-19 pandemic. It was conducted retrospectively by analyzing SARS-CoV-2 whole genomes of ∼30 kb. These genomes were isolated from human infections - asymptomatic and symptomatic. From December 2019 to September 2020, a total of 5,959 complete genomes with their associated clinical and patient data were retrieved from the Global Initiative on Sharing Avian Influenza Data (GISAID) database [13]. The demographic data included age (grouped into four categories: 0 – 19, 20 – 39, 40 – 59 and ≥ 60 years), gender and the geographical region (continent) where the infection was diagnosed. The associated metadata included clinical outcomes - asymptomatic (no symptoms) or symptomatic (mild or severe/critical). Other metadata included specimen type - upper respiratory tract (URT) or lower respiratory tract (LRT) and the technology/chemistry used to sequence the viral genome.

### Sequence alignments and selection of polymorphic loci

The genomes were aligned to the Wuhan reference strain (NC_045512.2) using minimap version 2.17, and the SNPs, including the corresponding amino acid changes, were called using the Geneious Prime SNP caller [14, 15]. Amino acids identical to the reference strain at the investigated loci were considered wild-type; else, they were considered mutants. Only polymorphic (≥ 2 alleles, i.e. amino acids, including the wild-type) loci with a minor allele frequency (MAF) of 0.01 were retained and analyzed in this study. These criteria were implemented to ensure unbiased construction of haplotypes (within a gene) and multilocus genotypes, i.e. MLGs (across ≥ two genes) [16]. An allele was designated by the amino acid followed by the codon (referred hereafter as locus) number. E.g. S D_614_ and S G_614_ indicate glutamine (wild-type allele) and glycine (mutant allele) at locus/codon 614 of the S protein.

### Population genetics

G**enetic diversity indices** – the number of haplotypes or MLGs, eMLGs (normalized MLG based on smallest sample) and expected heterozygosity (*H*_*e*_) were estimated using *poppr* V2.8.5 [16]. The eMLG was then plotted using the R package *Vegan* [17], as a rarefaction curve to estimate the depth/richness in sampling. The evenness (E5) statistic was used to evaluate whether the haplotypes or eMLGs found within the population were evenly distributed. Its score ranges from 0 (presence of predominant haplotypes or MLGs) to 1 (haplotypes or MLGs are evenly distributed). The *H*_*e*_ is a measure of genetic diversity, scoring from 0 (no genetic diversity, i.e., genomes carry the same haplotype or MLG) to 1 (complete diversity, i.e., genomes carry unique haplotypes or MLGs).

To determine **linkage disequilibrium (LD)**, i.e., non-random association of alleles at two or more loci, the standardized index of association 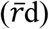 was estimated using *poppr*. The presence of genomes with identical MLGs, i.e. clones within a population, can overestimate the LD [18]. To account for this, the LD was clone-corrected using the dataset consisting of unique MLGs. To determine whether the LD was ‘structured’ between specific gene pairs, a pairwise LD was performed as described elsewhere [19]. The 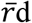 score ranges from 0 to 1, with 0 indicating no LD and 1 indicating complete LD. The statistical significance of the score was supported by a P-value < 0.05.

### Genetic differentiation

(Nei’s GST) among MLG populations within and between continents was estimated using *mmod* [20]. The G_ST_ score ranges from 0 (no genetic differentiation, i.e., populations are similar or have identical MLGs) to 1 (i.e., complete genetic differentiation, i.e., populations are dissimilar or have unique MLGs); values ranging from 0 to 0.09, 0.1 to 0.19 and ≥ 0.2 indicate little, moderate and great genetic differentiation, respectively [21]. We then performed a **Discriminant Analysis of Principal Components (DAPC)** – a multivariate method for identifying **genetic clusters** of closely related MLGs [22]. Briefly, the global MLG dataset was trained on a K-means algorithm, implemented in *adegenet* [22] to identify the optimum number of genetic clusters within the global population. The DAPC was then performed on the genetic clusters retained during a PCA by maximizing the genetic variance between populations while minimizing the variance within populations [23]. The DAPC assigned population membership probability to each MLG, which was plotted using *ggplot2* [24]. To visualize the genetic relationships and clonal complexes among the MLGs, the goeBURST FULL MST algorithm implemented in *Phyloviz* V2 was used to construct networks of minimum-spanning trees [25].

### Statistical analysis

Statistical analysis was performed in R v3.5.2 [26] and STATA v16 [27]. Proportions were compared using the chi-square or Fisher’s exact test. Multiple testing was adjusted using the Holm-Bonferroni method. Logistic regression was performed to determine the association between the study variables and the odds of harbouring a mutant allele. Age, geographical region, and gender were considered possible confounders and adjusted for in the final model. The adjusted odds ratio (OR) was considered statistically significant for all analysis where the P-value was < 0.05.

## Results and discussion

### Demographics of the study population

The majority of the SARS-CoV-2 whole genomes we sampled (N=5,959) from GISAID between December 2019 and September 2020 were reportedly from Asia (26.5%), Europe (19.9%) and Oceania (27.6%), with ≤ 10% each being from Africa, North America and South America (Table 1). Despite significant differences (P-value ≤ 0.005) in the proportion of genomes sampled for the study variables, > 35.0% of the genomes sampled in Africa and Oceania were reportedly from the 20 -39 years age group. In contrast, the majority of genomes (> 31.0%) from the rest of the world were reportedly from the 40 – 59 years age group (Table 1, S1 and S2). These data are consistent with the age disparities in COVID-19 cases [28, 29]. Except in Africa, where a significantly higher proportion of genomes were isolated from females (56.2%, P-value = 0.002), most were reportedly isolated from males.

**Table 1.** Demographics of the study population

| Factor | Category | Global<br>5959 | Africa<br>601 | Asia<br>1579 | Europe<br>1188 | N.America<br>597 | Oceania<br>1646 | S.America<br>348 |
| --- | --- | --- | --- | --- | --- | --- | --- | --- |
| **Age group#**<br>(years) | 0 – 19 | 323 (5.4) | 68 (11.3) | 123 (7.8) | 43 (3.6) | 20 (3.6) | 58 (3.5) | 11 (3.2) |
|  | 20 – 39 | 2054 (34.5) | 276 (45.9) | 559 (35.4) | 273 (22.9) | 196 (32.8) | 634 (38.5) | 116 (33.3) |
|  | 40 – 59 | 1996 (33.5) | 185 (30.8) | 561 (35.5) | 389 (32.7) | 217 (36.4) | 515 (31.3) | 129 (37.1) |
|  | 60+ | 1586 (26.6) | 72 (11.9) | 336 (21.3) | 483 (40.7) | 164 (27.5) | 439 (26.7) | 92 (26.4) |
| **Gender#** | Females | 2664 (44.7) | 338 (56.2) | 581 (36.8) | 581 (48.9) | 250 (41.9) | 754 (45.8) | 160 (45.9) |
|  | Males | 3295 (55.3) | 263 (43.8) | 998 (63.2) | 607 (51.1) | 347 (58.1) | 892 (54.2) | 188 (54.0) |
| **Clinical#**<br>**Severity** | Asymptomatic | 60 (1.5) | 0 (0.0) | 41 (2.7) | 18 (1.9) | 0 (0.0) | 0 (0.0) | 1 (0.4) |
|  | Mild | 557 (14.1) | 15 (2.5) | 134 (8.8) | 64 (6.7) | 282 (47.2) | 2 (12.5) | 60 (24.9) |
|  | Severe | 3321<br>(84.33) | 586 (97.5) | 1356(88.6) | 870 (91.4) | 315 (52.8) | 14 (87.5) | 180 (74.7) |
|  | Missing data* | 2021 | 0 | 48 | 236 | 0 | 1630 | 107 |

### The majority of the SARS-CoV-2 genomes were reportedly isolated from throat swabs and were sequenced using Illumina

Half of the genomes we analyzed had the associated data on the specimen type collected for diagnosis or isolating the virus genome. URT specimens constituted 92.0%. Of these, throat swabs were the majority (65.1%) (Figure S2). This was observed for all the study variables (Figure S3-S5) except in South America, where more than 60.3% of the genomes were isolated from nose swabs (Figure S4). Globally, more than 68.0% of the genomes we sampled were reportedly sequenced using Illumina except in Asia and South America, where a higher proportion of genomes were reportedly sequenced using Ion Torrent (39.53%) and Nanopore (43.10%), respectively (Figure S6).

### Seventy-four polymorphic loci were selected for multilocus genotyping of SARS-CoV-2 genomes

The majority of mutations in SARS-CoV-2 variants have been considered neutral, i.e. associated with demographic processes [30, 31]. A few others, considered homoplasic (recur independently) and adaptive (associated with viral transmissibility and/or pathogenicity), have been detected in clinical infections circulating worldwide [9, 31, 32]. Based on these reports and our filtering criteria of a MAF ≥ 0.01, 74 polymorphic loci (≥ 2 alleles) were used to construct the MLGs (Figure S7). These loci are located within ORF1ab – NSP1, NSP2, NSP3, NSP4, NSP5, NSP8, NSP12, NSP13 and NSP14, two accessory proteins - ORF3a and ORF8, and three structural proteins - M, N and S (Table S3). The moderate genetic differentiation (Gst = ∼0.10) observed for these loci demonstrate their utility as markers for differentiating SARS-CoV-2 variants (Table S4). The *orf1ab* gene was the most polymorphic loci (Table S3-S4), indicating a mutational hotspot in SARS-CoV-2 [33]. However, its low *H*_*e*_, ≤ 0.44, compared to the moderate-to-high *H*_*e*_, ≥ 0.5, in the *orf3a*, S and N genes is consistent with previous reports using nucleotide data [34]. Furthermore, our data also suggest that most mutations in the *orf1ab* were not under strong selection compared to those in the accessory and structural genes [30, 31, 35]. Indeed, B cell epitopes on the N and S proteins were shown to be highly diversified, allowing the virus to evade host immune responses [36].

### The spatiotemporal selection of variants carrying mutant alleles - ORF1ab L_4715,_ S G_614_, and N K_203_ and R_204_ was associated with spikes in COVID-19 cases

Of the 149 mutant alleles detected among the 74 loci (Figure S8-S10 and Table S4), eight alleles (Figure 1) were considered putatively adaptive, having been previously associated with infectivity, pathogenesis and host immune dysregulation [4]. The first four - ORF1ab L_4715_ (located in NSP12/RdRP), S G_614_, and N K_203_ and R_204_ rose sharply in frequency from < 6% in February to > 90% in August. This rise was associated with significant peaks in global COVID-19 cases between March and September (Figure 1) [37]. The remaining four alleles, including ORF1ab I_265_ (NSP1), ORF1ab F_3606_ (NSP5), ORF8 S_84_ and ORF3a H_57,_ were detected ‘transiently’ at a lower frequency, ranging from 4 to 35% (Figure 1 and Figure S8-S10). In contrast, most of the remaining 141 alleles circulated at a lower frequency, < 25%, and were not detected throughout the study period and in all continents (Figure S8-S10 and Table S4). These alleles, particularly those in the ORF1ab, have been considered neutral [31]. It is worth noting that the N501Y mutation in the S protein, associated with higher infectivity among UK and South African variants [5, 6], was carried by 0.8% of the genomes we sampled from Australia, Oceania. These genomes were reportedly isolated in June, suggesting that the S Y_501_ allele emerged earlier than previously reported [38].

**Figure 1.**
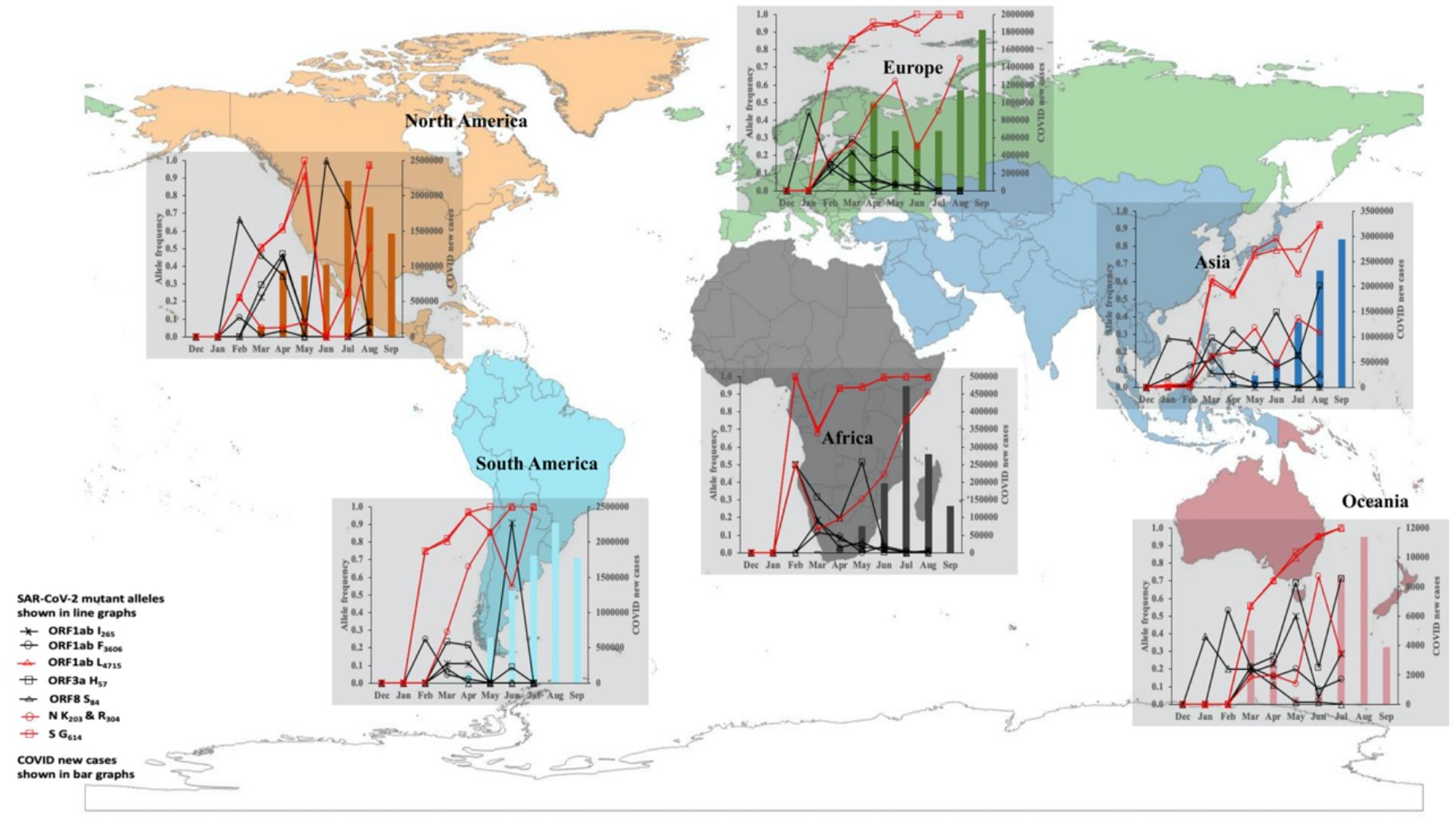
Spatiotemporal prevalence of eight SARS-CoV-2 mutant alleles and COVID-19 new cases. The prevalence data for the alleles and newly confirmed cases (WHO report 2020) are reported for December 2019 to September 2020. Four mutant alleles - ORF1ab L_4715_, S G_614_, and N K_203_ and R_204_ were associated with spikes in COVID-19 cases.

### The risk of harbouring a mutant allele varied with age, gender, geographical region and COVID-19 phenotype

A critical gap in the surveillance of SARS-CoV-2 infections and understanding COVID-19 clinical severity is the lack of data on the relationship between SARS-CoV-2 variants causing clinical infections and the clinical and demographic factors of infected individuals. To investigate this, we estimated the risk of harbouring a wild-type versus mutant allele at eight informative loci among our study variables. These informative loci – ORF1ab (265, 3606 and 4715), ORF3a 57, ORF8 84, N (203 and 204) and S 614 had alleles that were evenly distributed (E.5 score ≥ 0.6) in the global population and recurred throughout the study period (Figure 1 and Table S3).

Briefly, the risk of harbouring a mutant allele was associated with age, gender and COVID-19 phenotype in all the six continents (Figure 2). Compared to patients below 20 years, patients aged 20+ years had a higher risk of harbouring mutant alleles at four loci - N 203 and 204 (Asian males), ORF3a 57 (Europe) and ORF8 84 (North America) (Figure 2). This risk among the 20+ years cohort was maintained at the S 614 locus for patients in Oceania but not in North America. Host immune responses to the accessory and structural proteins have been implicated as drivers of SARS-CoV-2 evolution, with immune responses of older patients associated with infections carrying mutant alleles [39]. Interestingly, compared to asymptomatic cases, severe cases in Africa, Asia and North America were more likely to harbour the S D_614_ allele (Figure 2). Although the S G_614_ has been associated with enhanced viral transmission, our data indicates that it was not associated with severe COVID-19 as reported elsewhere [40].

**Figure 2.**
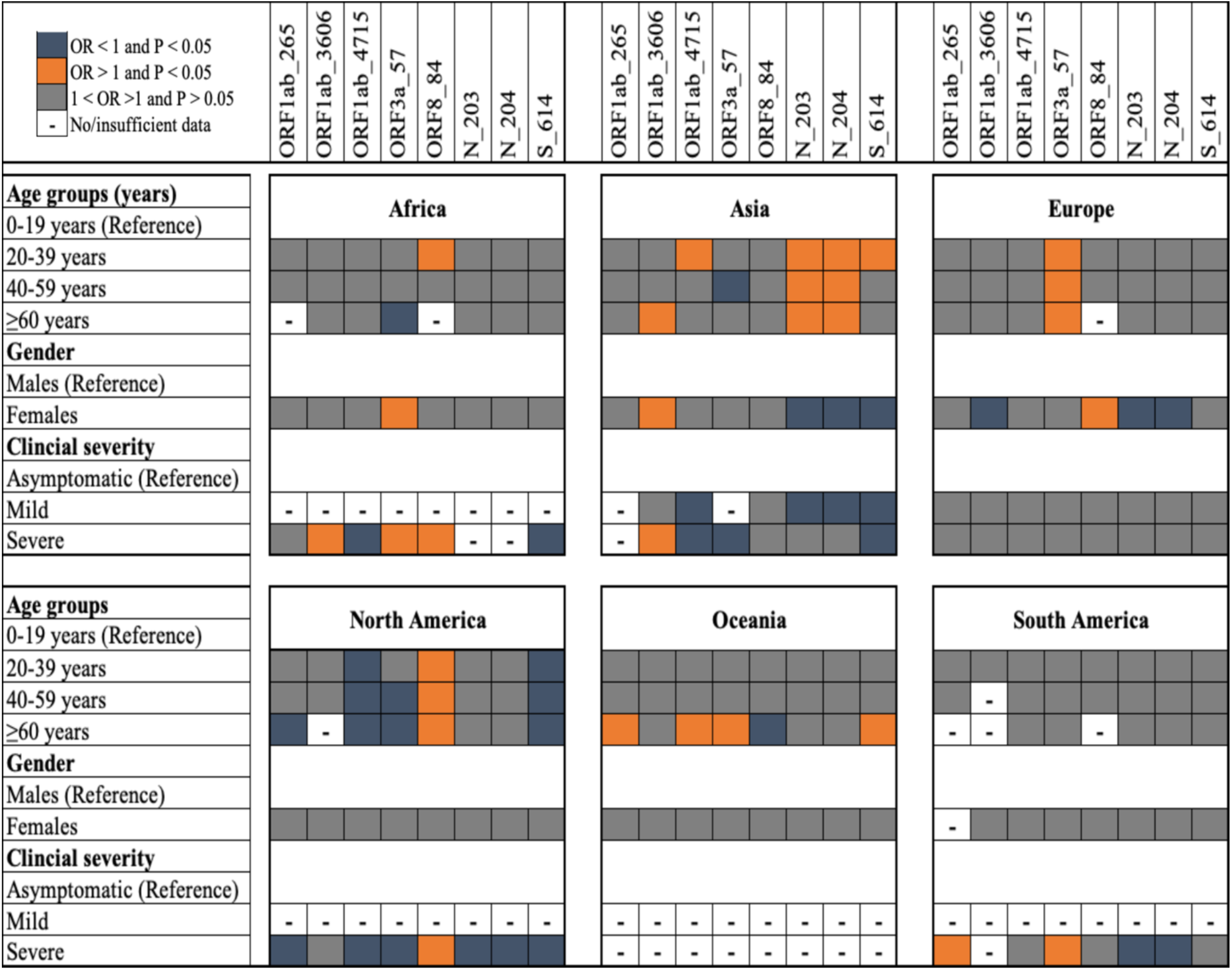
Eight informatic loci in SARS-CoV-2 associated with carriage of mutant alleles among different age groups, gender, geographical regions and COVID-19 clinical phenotype. The adjusted odds ratio (OR) with the P-value are shown with orange, blue and grey matrices indicating OR > 1 and P-value < 0.05, OR < 1 and P-value < 0.05 and 1< OR > 1 and P-value > 0.05, respectively. The OR was not estimated for study variables with < 5 samples.

### The majority of SARS-CoV-2 MLGs within each continent were detected except in Asia and Oceania

A major hurdle to tracking the spread and understanding the evolution of SARS-CoV-2 is the complexity and close relatedness of infecting genomes within and between different geographical regions. Therefore, we utilized the 74 polymorphic loci to differentiate the 5,959 genomes by constructing MLGs. We then used the MLG data to measure the richness in our sampling and to interrogate the population genetics of SARS-CoV-2 at the global and continental levels.

The MLGs were moderately distributed, as indicated by the E.5 score ranging from 0.41 for Europe to 0.60 for South America. Asia and Oceania had the highest number of observed (≥ 181) and expected (≥ 77) unique MLGs (Table S5). South America had the lowest number, 40 (Table 3 and S6). The plateauing of the rarefaction curve for Africa, Europe and the Americas indicated that no new MLGs were detected with further sampling in these populations (Figure 3A). In contrast, the steep rise in Asia and Oceania’s curve suggested that we had under-sampled and therefore did not capture most of the MLGs within these populations. As indicated by the positive correlation (r = 0.94, P-value = 0.017) between sample size and MLG abundance, deep sampling is critical for detecting most SARS-CoV-2 variants causing COVID-19 in affected communities. However, detecting most variants may be challenging in situations where logistics for COVID-19 testing are inadequate. Indeed, < 2% of the genomes we sampled were reportedly isolated from patients with asymptomatic infections. Considering that asymptomatic infections constitute ∼80% of all COVID-19 cases and refuel and sustain the virus’s transmission worldwide [41], they must be included during surveillance.

**Figure 3.**
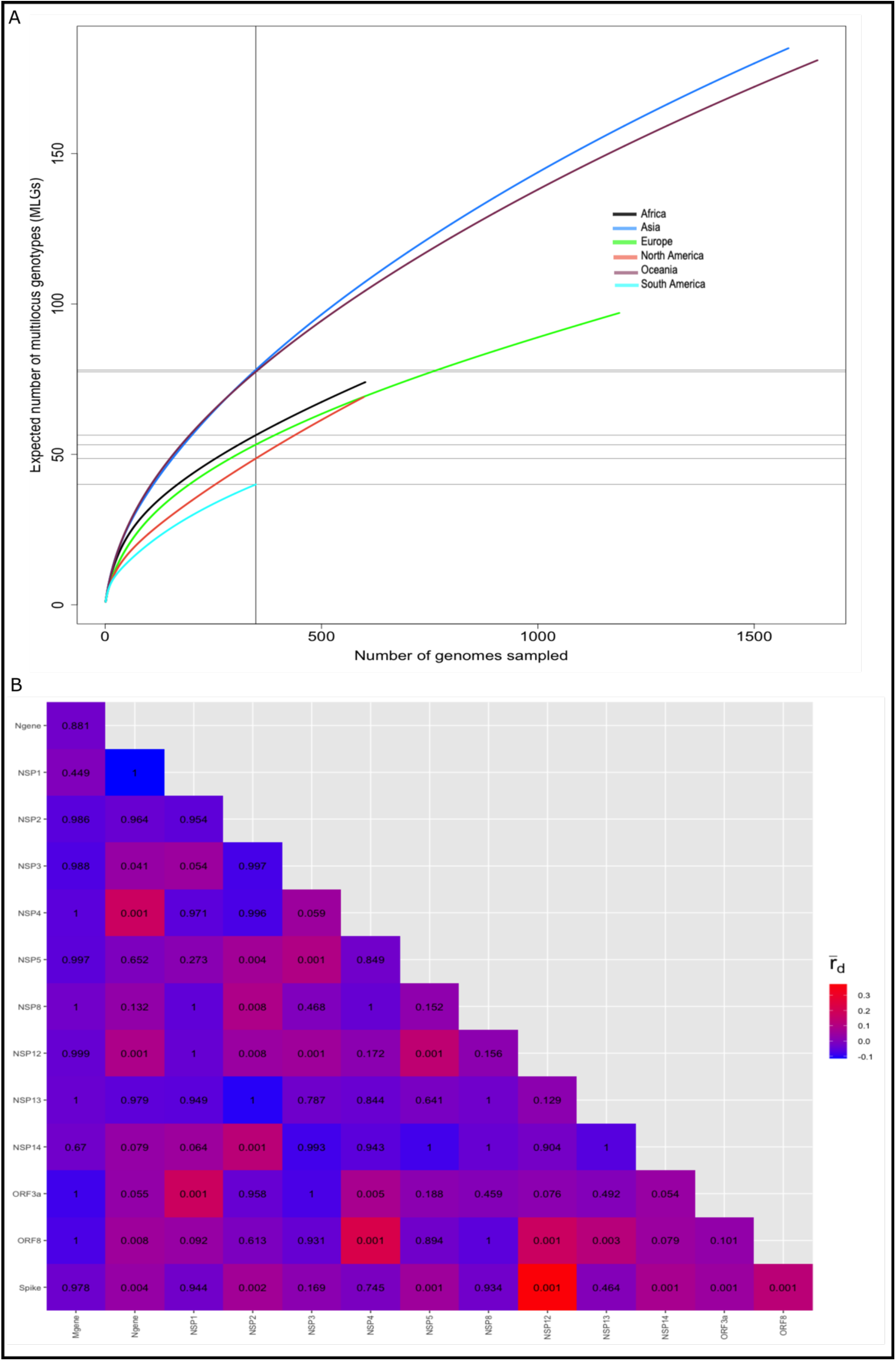
Richness in MLG sampling and the standardized index of association 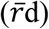 among genes in the global SAR-CoV-2 population sampled from December 2019 to September 2020. A. Rarefaction curve of the MLGs sampled in each geographical region. The plateau in the curves indicated no new MLGs were detected with further sampling in Africa, Europe, the American SARS-CoV-2 populations. B. Pairwise LD estimates among genes in the 5,959 genomes sampled globally. The 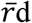 ranges from 0 (no LD) to 1 (complete LD). The values in the coloured heatmap indicate the P-value associated with the pairwise 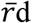 estimates. The strongest LD signal (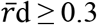, P-value < 0.001) was detected between NSP12 and S, NSP1 and ORF3a, NSP4 and ORF8, and NSP4 and N.

### Linkage structures in SARS-CoV-2 populations vary among geographical regions

We investigated the possibility that specific alleles could be linked and contribute to SARS-CoV-2 fitness, including transmissibility and pathogenicity. We first quantified the genetic diversity of SARS-CoV-2 populations in the different geographical regions. The multilocus genetic diversity was lowest in South America (*H*_*e*_ = 0.15) and highest in Asia, North America and Oceania (*H*_*e*_ ≥ 0.26) (Table 2). Our data is consistent with previous reports indicating that SARS-CoV-2 phylogenies in the latter two regions depicted the diversity that existed worldwide as of July 2020 [37, 42, 43]. There was significant genome-wide LD (non-random association among alleles) both at the global and continental levels (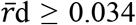, P-value < 0.001) (Table 2). However, the decay of this LD (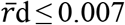, P-value ≥ 0.166) in Africa, Europe and South America after repeating the analysis with the unique MLGs was indicative of clonal SARS-CoV-2 transmission in these geographical regions. Indeed, multiple outbreaks in Europe and South America were due to local transmissions and were largely associated with clusters of closely related infections [43, 44].

**Table 2:** Genetic diversity estimates for SAR-COV-2 populations circulating globally.

| Continent | N | MLGs | eMLGs (SE) | E.5 | $H_e$ | $\bar{r}d$ (P-value) | $\bar{r}d$-cc (P-value) |
| --- | --- | --- | --- | --- | --- | --- | --- |
| Africa | 601 | 74 | 56.4 (3.1) | 0.57 | 0.19 | 0.034 (0.001) | 0.007 (0.230) |
| Asia | 1579 | 185 | 78.1(5.2) | 0.48 | 0.26 | 0.080 (0.001) | 0.022 (0.001) |
| Europe | 1188 | 97 | 53.2 (3.8) | 0.41 | 0.18 | 0.082 (0.001) | 0.009 (0.166) |
| N.America | 597 | 69 | 48.6 (3.3) | 0.47 | 0.30 | 0.219 (0.001) | 0.065 (0.001) |
| Oceania | 1646 | 181 | 77.5 (5.0) | 0.45 | 0.30 | 0.082 (0.001) | 0.020 (0.001) |
| S.America | 348 | 40 | 40.0 (0.0) | 0.60 | 0.15 | 0.080 (0.001) | 0.007 (0.278) |
| Total | 5959 | 472 | 95.3 (5.8) | 0.34 | 0.26 | 0.079 (0.001) | 0.019 (0.001) |

Interestingly, we observed that the genome-wide LD was driven by specific gene pairs, i.e., ‘structure’. We detected the strongest LD signal (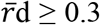, P-value < 0.001) between NSP12 and S, NSP1 and ORF3a, NSP4 and ORF8, and NSP4 and N (Figure 3B and Figure S12), consistent with previous reports using nucleotide data [39]. While the former three LD structures either decayed or were maintained at the continental level, the NSP12 and S LD structures were consistently prominent in all the geographical regions (Figure S11). A probable driver of the NSP12-S LD structure could be the strong co-selection of the *orf1ab* L_4715_ and S G_614_ alleles, which putatively enhance viral replication and infectivity, respectively [45]. Our findings imply that the evolutionary pressures shaping the virus vary in different geographical regions - with public health interventions, demographic factors, and host immune responses being the most likely key drivers [39, 46, 47]. Nonetheless, it is worth noting that these LD structures may change as the virus continues to evolve, underscoring the need for continuous genomic surveillance.

The number of observed MLGs was normalized by the smallest sample size to obtained the expected MLGs (eMLGs) with the standard error (SE). The standardized index of association 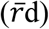 was clone-corrected 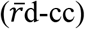 using the unique MLGs dataset. N. America (North America) and S. America (South America).

### SARS-CoV-2 MLGs in Asia and Oceania were representative of the global MLG population

The minimum spanning tree (Figure 4A) was drawn to visualize the network relationships among the 472 unique MLGs. Here, at least 11 major clusters, including eight global clusters (GC1 to 8), were identified. Nearly all clusters comprised a considerable number of Asian MLGs (Figure 4A), indicative of admixture populations, as shown in Figure 4B. The majority of MLGs in each continent were predicted to have 20-50% geographical assignment to Asia and Oceania (Figure 4B). This data suggests that the majority of SARS-CoV-2 variants in the world are of Asian descent. It also supports contact tracing data that showed that most SARS-CoV-2 cases during the early (March to May) stages of the pandemic were linked to imported cases from Asia [48]. However, a few MLGs were unique to each continent, as indicated by the 40-60% within-continent membership assignments. Prominent among continental clusters were the AC1 and OC1 clusters, detected in Asia and Oceania, respectively (Figure 4A). The ∼60% membership assignment of African MLGs to Europe (Figure 4B) suggested that most African infections were more likely to have been imported from Europe, contradicting previous reports of an American source based solely on travel data [49, 50]. This underscores the need to build strong surveillance systems utilizing both travel and genomic data.

**Figure 4.**
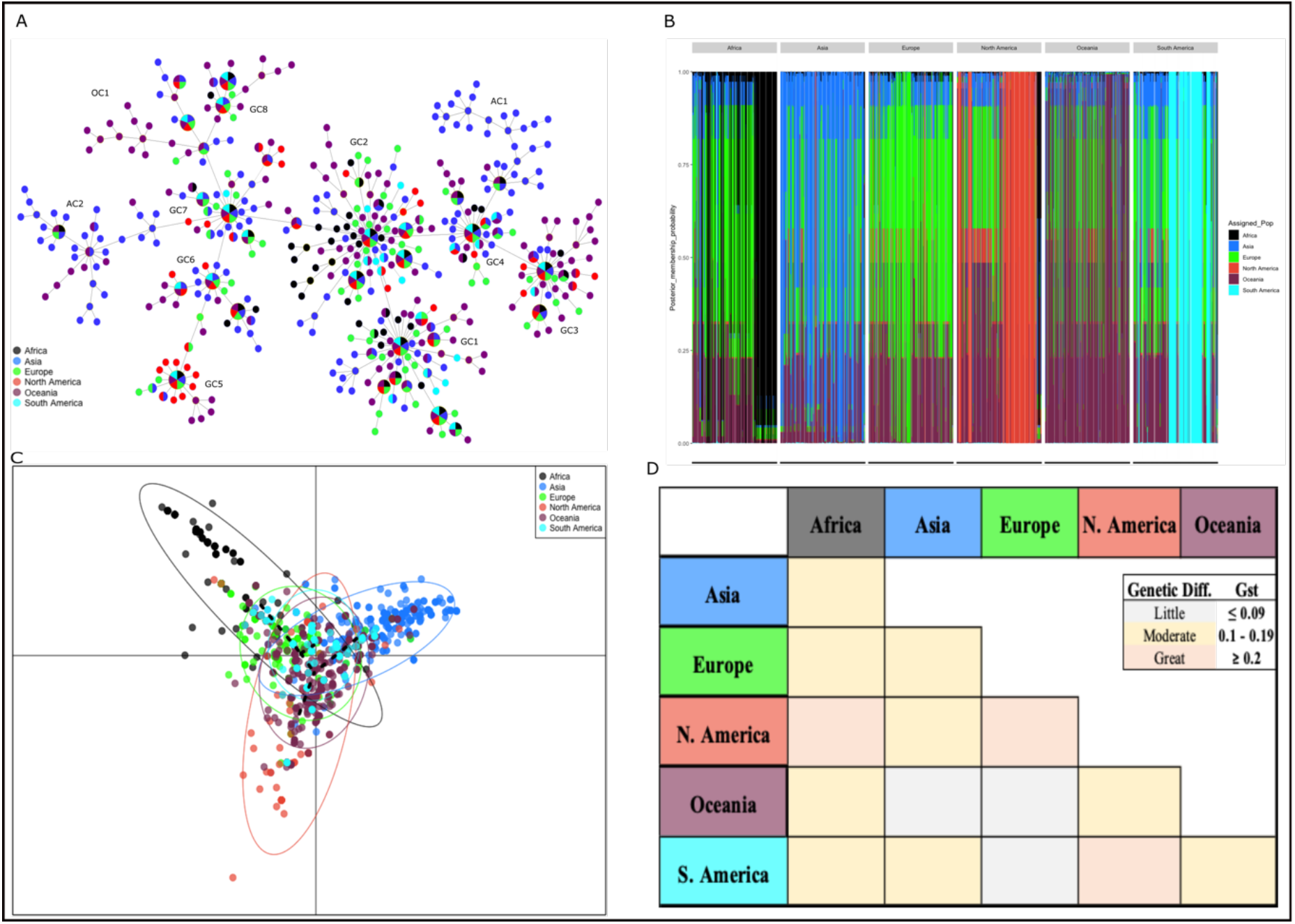
Genetic relatedness and differentiation among SAR-CoV-2 MLGs from different geographical regions. A. Relatedness among the 472 unique MLGs in the global population. Eleven clusters including eight global clusters (GC1-8) and three continental clusters – Asia (AC1-2) and Oceania (OC1) were detected. Nearly all clusters contained a considerable number of Asian MLGs. B. Population membership assignment of each MLG. Admixture populations were prominent in all geographical regions. C. DAPC analysis identified one global cluster (MLGs from all regions, central axis of PCA plot) and three continental clusters – Africa, Asia and North America. D. Moderate to high genetic differentiation (Nei’s Gst) was observed among MLGs from different continents.

### SARS-CoV-2 MLGs were genetically differentiated within and between continents

We detected many mutant alleles restricted to each continent (Table S6), suggesting geographical structuring in SARS-CoV-2 populations. Asia had the highest number of unique (?) alleles, 19 in total, including ORF1ab D_1812_, P_676_, G_2586_ and I_6297,_ being detected in 7.4, 4.5, 3.1 and 2.8%, respectively of genomes sampled. Among the 16 private alleles detected in Oceania, ORF1ab S_271_ (4.4%) was predominant. Europe had 12 private alleles with S G_320_ (10.7%) being predominant, while ORF3a S_251_ (5.4%) was predominant among eight private alleles detected in North America. Nine private alleles, including ORF1ab H_4080_ (15.6%) and N L_187_ (8.5%), were detected in Africa. Only two private alleles – ORF3a A_165_ (0.6%) and ORF8 A_62_ (0.6%) were detected in South America.

Further support for the geographical structuring in the global SARS-CoV-2 populations was obtained from the DAPC analysis in which we identified four main genetic clusters (Figure 4C). One cluster consisted of a subset of MLGs from each continent, consistent with the previously described admixture populations in Figure 4B. In contrast, the other three clusters consisted of MLGs, predominantly from Africa, Asia and North America (Figure 4C). A minor proportion of MLGs from Oceania showed clinal differentiation into North America (Figure 4B and C), which likely represent closely related SARS-CoV-2 variants that spread between the two regions [51]. A minor proportion of European and South American MLGs showed clinal differentiation into the African cluster (Figure 4C). The Gst estimates also supported evidence of geographical structuring. There was moderate-to-high genetic differentiation (Gst ≥ 0.204) among the continents except between Oceania, separately with Asia and Europe, and between Europe and South America, where there was little genetic differentiation (Gst ≤ 0.079) (Figure 4D).

Spatial connectivity, including cross-border migrations and international travel, is a major conduit for spreading the virus (i.e., resulting in gene flow) among countries. Hence, we expected to see little genetic differentiation among ‘regional blocks’ within a continent. This hypothesis was valid for Europe, where there was little to moderate genetic differentiation (Gst ≤ 0.181) among regional blocks except between Eastern and Northern Europe (Gst = 0.248) (Figure S12). Interestingly, we detected moderate-to-high genetic differentiation (Gst ≥ 0.111) among regional blocks in Africa and Asia (Figure S12). This may be a reflection of the fast and strict travel bans that were put into effect early during the spread of the virus in both regions.

## Conclusion

The disproportionate distribution of SARS-CoV-2 genomes among the young and older age groups in this study was representative of the age distribution of COVID-19 cases reported globally. Throat swabs were the preferred specimen for COVID-19 diagnosis, but the invasive nature involved in sampling may limit its utility for surveillance. In building a robust and efficient surveillance system, access to affordable sequencing and rapid analysis of complex genomic data will be seminal to inform control efforts. The utility of the 74 polymorphic loci as markers for studying the epidemiology and population genetics of SARS-CoV-2 infections represents significant progress in developing molecular tools for SARS-CoV-2 Surveillance. In particular, the eight informatic loci revealed contrasting epidemiology and transmission dynamics of the virus among different demographics, geographical regions and COVID-19 phenotypes. The selection of alleles at these loci and the maintenance of key LD structures in the infecting genomes indicate that the virus is evolving and adapting resulting in enhanced transmissibility to humans. An effect of this evolution was the structuring we observed in the viral population, which allowed us to differentiate closely related variants between different geographical regions genetically. Future studies can include additional loci to increase the differentiation among variants within the same geographical region.

## Limitations

The data presented in this study needs further investigations to draw definite conclusions on the association between age, gender and geographical region and the SARS-CoV-2 variant causing COVID-19. Nearly all the genomes we sampled did not have metadata on where the patient got infected besides the country where the infection was diagnosed and/or genome isolated. Thus, it was difficult to infer a source of infection based solely on the MLG data.

## Data Availability

All relevant data are contained in the manuscript and the supplementary data

## Author Contributions

CAN, RA and JSR conceived and designed the study. CAN and FC acquired the data and performed the analysis. CAN and FC drafted the manuscript. RA and JSR critically revised the manuscript. All authors read and approved the manuscript for publication.

## Acknowledgements

We are grateful to the authors from the laboratories responsible for obtaining the specimens and the submitting laboratories where genetic sequence data were generated and shared via the GISAID Initiative, on which this research is based.

