## Supplementary data for "Contrasting epidemiology and population genetics of COVID-19 infections defined with 74 polymorphic loci in SARS-CoV-2 genomes sampled globally"

Table S1. Pairwise t-test (Holms Corrected) for difference in the proportion of genomes sampled among the four age groups in the different geographical regions.

| | Africa | Asia | Europe | North America | Oceania |
|---|---|---|---|---|---|
| Asia | 0.0001 | - | - | - | - |
| Europe | 0.0001 | 0.0001 | - | - | - |
| North America | 0.0001 | 0.00015 | 0.0001 | - | - |
| Oceania | 0.0001 | 0.00214 | 0.0001 | 0.3015 | - |
| South America | 0.0001 | 0.00559 | 0.0001 | 0.84368 | 0.53861 |

P-value < 0.05 was considered statistically significant.

Table S2. Pairwise t-test (Holms Corrected) for difference in the proportion of genomes sampled among the genders in the different geographical regions.

| | Africa | Asia | Europe | North America | Oceania |
|---|---|---|---|---|---|
| Asia | 0.0001 | - | - | - | - |
| Europe | 0.0242 | 0.0001 | - | - | - |
| North America | 0.0001 | 0.1939 | 0.032 | - | - |
| Oceania | 0.0001 | 0.0001 | 0.4786 | 0.4786 | - |
| South America | 0.0184 | 0.017 | 0.6614 | 0.655 | 0.9538 |

P-value < 0.05 was considered statistically significant.

### Table S3: The 74 loci/codons considered polymorphic and included in the study.

| Genetic diversity in genes coding for non-structural proteins | | | | | |
|---|---|---|---|---|---|
| Protein | Gene | Codon | No of alleles | He | E.5 |
| NSP1 | ORF1ab | 75 | 2 | 0.02 | 0.36 |
| | ORF1ab | 207 | 3 | 0.01 | 0.32 |
| | ORF1ab | 265 | 3 | 0.24 | 0.63 |
| NSP2 | ORF1ab | 271 | 3 | 0.02 | 0.35 |
| | ORF1ab | 300 | 2 | 0.02 | 0.36 |
| | ORF1ab | 378 | 2 | 0.05 | 0.42 |
| | ORF1ab | 392 | 3 | 0.01 | 0.31 |
| | ORF1ab | 454 | 2 | 0.01 | 0.33 |
| | ORF1ab | 481 | 2 | 0.01 | 0.33 |
| | ORF1ab | 676 | 3 | 0.02 | 0.36 |
| | ORF1ab | 739 | 2 | 0.09 | 0.47 |
| | ORF1ab | 765 | 2 | 0.02 | 0.35 |
| | ORF1ab | 876 | 4 | 0.01 | 0.30 |
| NSP3 | ORF1ab | 944 | 2 | 0.01 | 0.32 |
| | ORF1ab | 971 | 4 | 0.03 | 0.34 |
| | ORF1ab | 972 | 2 | 0.00 | 0.21 |
| | ORF1ab | 1036 | 4 | 0.02 | 0.33 |
| | ORF1ab | 1100 | 3 | 0.03 | 0.37 |
| | ORF1ab | 1246 | 2 | 0.03 | 0.38 |
| | ORF1ab | 1812 | 3 | 0.04 | 0.39 |
| | ORF1ab | 2015 | 4 | 0.04 | 0.39 |
| | ORF1ab | 2016 | 3 | 0.08 | 0.46 |
| | ORF1ab | 2129 | 3 | 0.03 | 0.37 |
| | ORF1ab | 2153 | 2 | 0.01 | 0.30 |
| | ORF1ab | 2202 | 4 | 0.01 | 0.31 |
| | ORF1ab | 2586 | 2 | 0.02 | 0.34 |
| | ORF1ab | 2702 | 2 | 0.02 | 0.34 |
| | ORF1ab | 2796 | 5 | 0.01 | 0.30 |
| NSP4 | ORF1ab | 3071 | 2 | 0.10 | 0.49 |
| | ORF1ab | 3278 | 2 | 0.03 | 0.38 |
| | ORF1ab | 3334 | 3 | 0.02 | 0.34 |
| NSP5 | ORF1ab | 3529 | 2 | 0.01 | 0.31 |
| | ORF1ab | 3606 | 3 | 0.21 | 0.60 |
| NSP8 | ORF1ab | 4080 | 2 | 0.03 | 0.38 |
| | ORF1ab | 4140 | 2 | 0.01 | 0.32 |
| NSP12 | ORF1ab | 4489 | 2 | 0.09 | 0.47 |
| | ORF1ab | 4715 | 4 | 0.42 | 0.85 |
| NSP13 | ORF1ab | 5809 | 3 | 0.01 | 0.33 |
| | ORF1ab | 5828 | 3 | 0.04 | 0.40 |
| | ORF1ab | 5865 | 3 | 0.04 | 0.40 |
| | ORF1ab | 5894 | 2 | 0.00 | 0.27 |
| NSP14 | ORF1ab | 6102 | 2 | 0.02 | 0.36 |
| | ORF1ab | 6158 | 2 | 0.02 | 0.35 |
| | ORF1ab | 6297 | 2 | 0.01 | 0.34 |
| | ORF1ab | 6302 | 2 | 0.00 | 0.21 |
| | ORF1ab | 6474 | 3 | 0.01 | 0.33 |

| Genetic diversity in genes coding for structural and accessory proteins | | | | | |
|---|---|---|---|---|---|
| Protein | Gene | Codon | No of alleles | He | E.5 |
| Membrane (M) | M | 3 | 3 | 0.03 | 0.36 |
| | M | 175 | 2 | 0.02 | 0.35 |
| Nucleocapsid (N) | N | 13 | 2 | 0.11 | 0.50 |
| | N | 22 | 4 | 0.01 | 0.29 |
| | N | 183 | 3 | 0.00 | 0.23 |
| | N | 187 | 2 | 0.02 | 0.34 |
| | N | 194 | 3 | 0.07 | 0.44 |
| | N | 197 | 3 | 0.10 | 0.48 |
| | N | 202 | 4 | 0.03 | 0.37 |
| | N | 203 | 5 | 0.39 | 0.80 |
| | N | 204 | 4 | 0.38 | 0.80 |
| | N | 292 | 3 | 0.03 | 0.38 |
| Spike glycoprotein (S) | S | 54 | 2 | 0.03 | 0.38 |
| | S | 320 | 5 | 0.04 | 0.39 |
| | S | 477 | 5 | 0.02 | 0.32 |
| | S | 485 | 2 | 0.01 | 0.31 |
| | S | 614 | 3 | 0.41 | 0.84 |
| | S | 1124 | 2 | 0.02 | 0.36 |
| | S | 1176 | 2 | 0.02 | 0.34 |
| ORF3a | ORF3a | 57 | 2 | 0.37 | 0.78 |
| | ORF3a | 99 | 3 | 0.01 | 0.31 |
| | ORF3a | 126 | 3 | 0.02 | 0.34 |
| | ORF3a | 165 | 3 | 0.01 | 0.31 |
| | ORF3a | 196 | 2 | 0.10 | 0.49 |
| | ORF3a | 251 | 5 | 0.09 | 0.43 |
| ORF8 | ORF8 | 24 | 4 | 0.05 | 0.40 |
| | ORF8 | 62 | 5 | 0.03 | 0.36 |
| | ORF8 | 84 | 3 | 0.21 | 0.60 |

Table S4. Genetic diversity within genes in SARS-CoV-2. The genetic diversity parameters are indicated by h (number of haplotypes), He (heterozygosity), evenness score (E.5) and genetic differentiation index (Gst).

| Gene | h | *He* | E.5 | Gst |
|---|---|---|---|---|
| NSP1 | 6 | 0.27 | 0.57 | 0.070 |
| NSP2 | 21 | 0.22 | 0.33 | 0.088 |
| NSP3 | 47 | 0.30 | 0.29 | 0.050 |
| NSP4 | 5 | 0.15 | 0.43 | 0.125 |
| NSP5 | 4 | 0.22 | 0.57 | 0.065 |
| NSP8 | 3 | 0.04 | 0.36 | 0.116 |
| NSP12 | 6 | 0.44 | 0.71 | 0.127 |
| NSP13 | 9 | 0.06 | 0.35 | 0.023 |
| NSP14 | 9 | 0.05 | 0.30 | 0.021 |
| ORF3a | 19 | 0.53 | 0.59 | 0.082 |
| ORF8 | 13 | 0.26 | 0.49 | 0.145 |
| Membrane | 4 | 0.05 | 0.35 | 0.015 |
| Nucleocapsid | 30 | 0.60 | 0.54 | 0.100 |
| Spike | 17 | 0.50 | 0.61 | 0.117 |
| Mean overall | 14 | 0.26 | 0.46 | 0.096 |

The genetic diversity parameters are indicated by h (number of haplotypes), He (heterozygosity), evenness score (E.5) and genetic differentiation index (Gst).

Table S5. Unique MLGs. Data attached in a separate excel file.

Attached in separate excel file.

### Table S6. Private alleles

| Gene | Gene | Codon/locus | Allele/mutation | Africa | Asia | Europe | North America | Oceania | South America |
|---|---|---|---|---|---|---|---|---|---|
| ORF1ab | NSP1 | 207 | R -> G | - | - | 0.1 | - | - | - |
| ORF1ab | NSP1 | 265 | T -> N | 0.2 | - | - | - | - | - |
| ORF1ab | NSP2 | 271 | P -> S | - | - | - | - | 4.4 | - |
| ORF1ab | NSP2 | 392 | G -> C | - | - | 0.2 | - | - | - |
| ORF1ab | NSP2 | 676 | Q -> P | - | 4.5 | - | - | - | - |
| ORF1ab | NSP2 | 676 | Q -> R | - | - | - | 0.2 | - | - |
| ORF1ab | NSP2 | 876 | A -> P | - | 0.1 | - | - | - | - |
| ORF1ab | NSP2 | 876 | A -> V | - | - | 0.1 | - | - | - |
| ORF1ab | NSP3 | 971 | P -> Q | - | 0.1 | - | - | - | - |
| ORF1ab | NSP3 | 972 | E -> V | - | 0.1 | - | - | - | - |
| ORF1ab | NSP3 | 1036 | D -> Y | 0.2 | - | - | - | - | - |
| ORF1ab | NSP3 | 1036 | D -> G | - | 0.1 | - | - | - | - |
| ORF1ab | NSP3 | 1100 | G -> A | - | - | - | - | 0.1 | - |
| ORF1ab | NSP3 | 1812 | A -> D | - | 7.4 | - | - | - | - |
| ORF1ab | NSP3 | 1812 | A -> V | - | - | - | - | 0.1 | - |
| ORF1ab | NSP3 | 2015 | S -> G | - | 0.1 | - | - | - | - |
| ORF1ab | NSP3 | 2015 | S -> N | - | 0.1 | - | - | - | - |
| ORF1ab | NSP3 | 2129 | A -> G | - | - | - | 0.2 | - | - |
| ORF1ab | NSP3 | 2153 | T -> I | - | - | - | - | 1.4 | - |
| ORF1ab | NSP3 | 2202 | T -> A | 0.2 | - | - | - | - | - |
| ORF1ab | NSP3 | 2202 | T -> N | - | 0.1 | - | - | - | - |
| ORF1ab | NSP3 | 2586 | V -> G | - | 3.1 | - | - | - | - |
| ORF1ab | NSP3 | 2796 | M -> T | - | 0.1 | - | - | - | - |
| ORF1ab | NSP4 | 3334 | G -> V | - | - | - | - | 0.1 | - |
| ORF1ab | NSP5 | 3529 | A -> V | - | - | - | - | 1.7 | - |
| ORF1ab | NSP5 | 3606 | L -> C | - | - | - | - | 0.1 | - |
| ORF1ab | NSP8 | 4080 | Y -> H | 15.6 | - | - | - | - | - |
| ORF1ab | NSP12 | 4715 | P -> T | - | - | 0.1 | - | - | - |
| ORF1ab | NSP12 | 4715 | P -> S | - | - | 0.1 | - | - | - |
| ORF1ab | NSP13 | 5809 | S -> P | - | - | - | - | 0.1 | - |
| ORF1ab | NSP13 | 5865 | Y -> F | - | - | - | 0.2 | - | - |
| ORF1ab | NSP14 | 6297 | T -> I | - | 2.8 | - | - | - | - |
| ORF1ab | NSP14 | 6302 | F -> L | - | 0.1 | - | - | - | - |
| ORF1ab | NSP14 | 6474 | V -> I | 0.2 | - | - | - | - | - |
| ORF3a | ORF3a | 99 | A -> S | - | 0.1 | - | - | - | - |
| ORF3a | ORF3a | 165 | S -> A | - | - | - | - | - | 0.6 |
| ORF3a | ORF3a | 251 | G -> D | 0.5 | - | - | - | - | - |
| ORF3a | ORF3a | 251 | G -> S | - | - | - | 5.4 | - | - |
| ORF3a | ORF3a | 251 | G -> C | - | - | - | - | 0.1 | - |
| ORF8 | ORF8 | 24 | S -> A | - | - | - | 0.2 | - | - |
| ORF8 | ORF8 | 24 | S -> P | - | - | - | 0.2 | - | - |
| ORF8 | ORF8 | 62 | V -> E | - | - | - | 0.2 | - | - |
| ORF8 | ORF8 | 62 | V -> F | - | - | - | - | 0.1 | - |
| ORF8 | ORF8 | 62 | V -> A | - | - | - | - | - | 0.6 |
| ORF8 | ORF8 | 84 | L -> F | - | 0.1 | - | - | - | - |
| Membrane | M | 3 | D -> Y | - | - | - | - | 0.1 | - |
| Nucleocapsid | N | 22 | D -> N | - | 0.1 | - | - | - | - |
| Nucleocapsid | N | 183 | S -> F | - | 0.1 | - | - | - | - |
| Nucleocapsid | N | 187 | S -> L | 8.5 | - | - | - | - | - |
| Nucleocapsid | N | 194 | S -> A | - | - | - | - | 0.1 | - |
| Nucleocapsid | N | 197 | S -> P | - | - | - | - | 0.1 | - |
| Nucleocapsid | N | 202 | S -> C | - | - | 0.1 | - | - | - |
| Nucleocapsid | N | 202 | S -> I | - | - | 0.1 | - | - | - |
| Nucleocapsid | N | 203 | R -> G | - | 0.1 | - | - | - | - |
| Nucleocapsid | N | 203 | R -> M | - | - | 0.1 | - | - | - |
| Nucleocapsid | N | 204 | G -> T | - | 0.1 | - | - | - | - |
| Nucleocapsid | N | 204 | G -> L | - | - | 0.1 | - | - | - |
| Nucleocapsid | N | 292 | I -> V | 0.2 | - | - | - | - | - |
| Spike | S | 320 | V -> G | - | - | 10.7 | - | - | - |
| Spike | S | 320 | V -> F | - | - | 0.1 | - | - | - |
| Spike | S | 320 | V -> I | - | - | - | 0.2 | - | - |
| Spike | S | 320 | V -> A | - | - | - | - | 0.1 | - |
| Spike | S | 477 | S -> R | 0.2 | - | - | - | - | - |
| Spike | S | 477 | S -> G | - | - | 0.1 | - | - | - |
| Spike | S | 485 | G -> R | - | - | - | - | 1.6 | - |
| Spike | S | 614 | D -> E | - | - | - | - | 0.1 | - |
| Total | | | 66 | 9 | 19 | 12 | 8 | 16 | 2 |

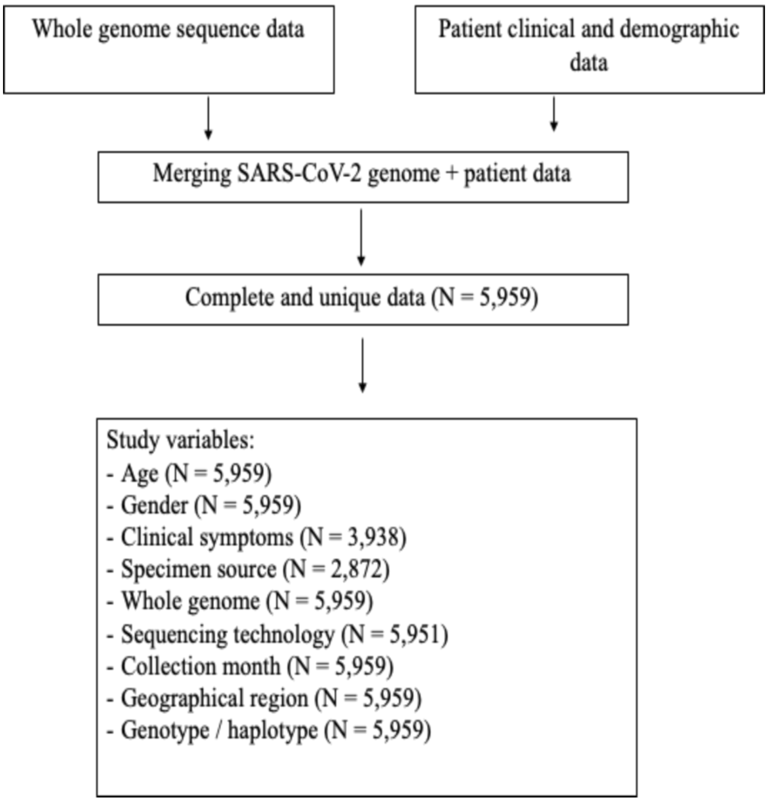


Figure S1. Workflow for data acquisition and cleaning prior to analysis.

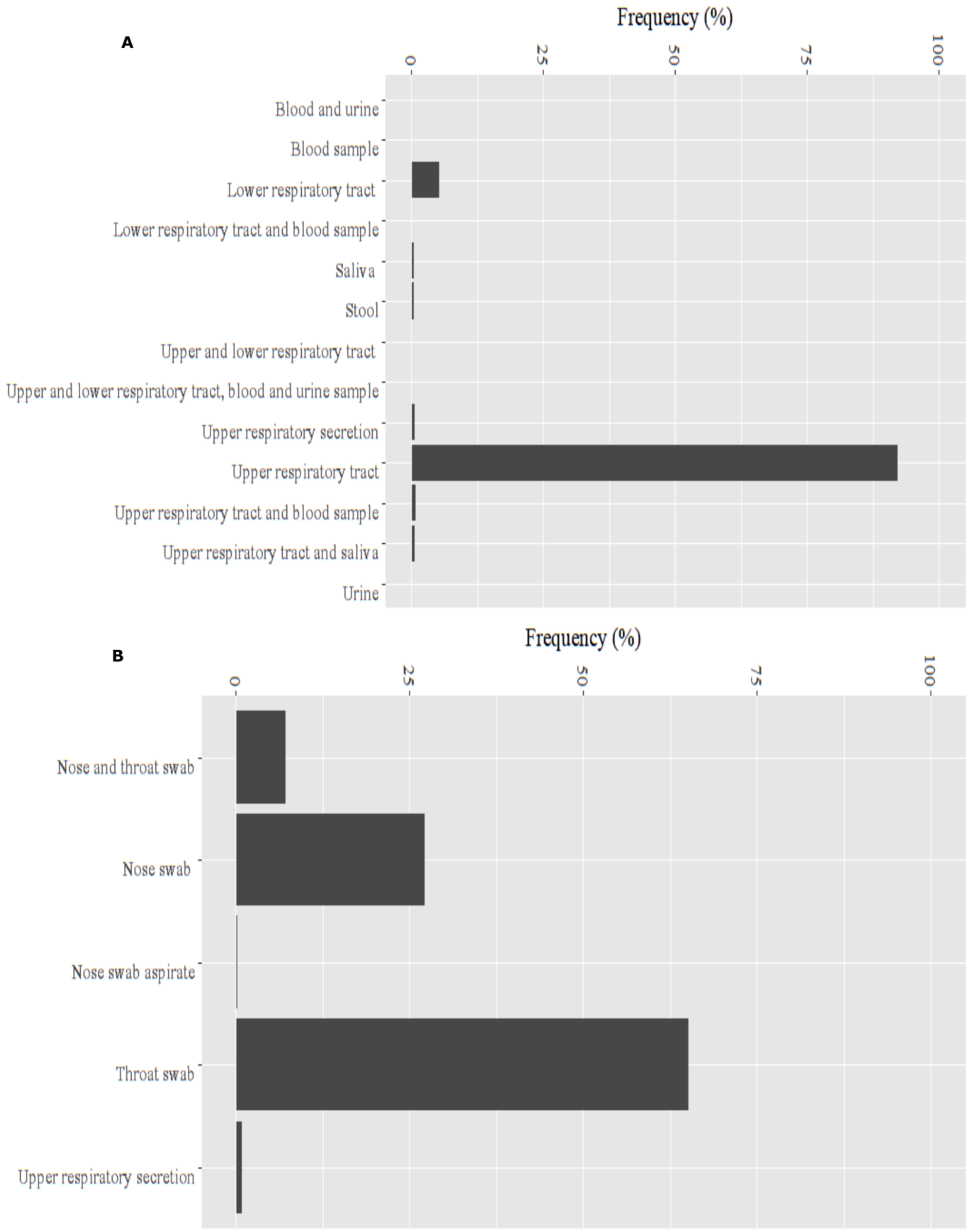


Figure S2. Specimen types collected for SARS-CoV-2 diagnosis and genome isolation. Upper graph details the proportion of genomes isolated using 13 specimen types. Lower graph details the proportion of genomes for the different upper respiratory tract (URT) sample types.

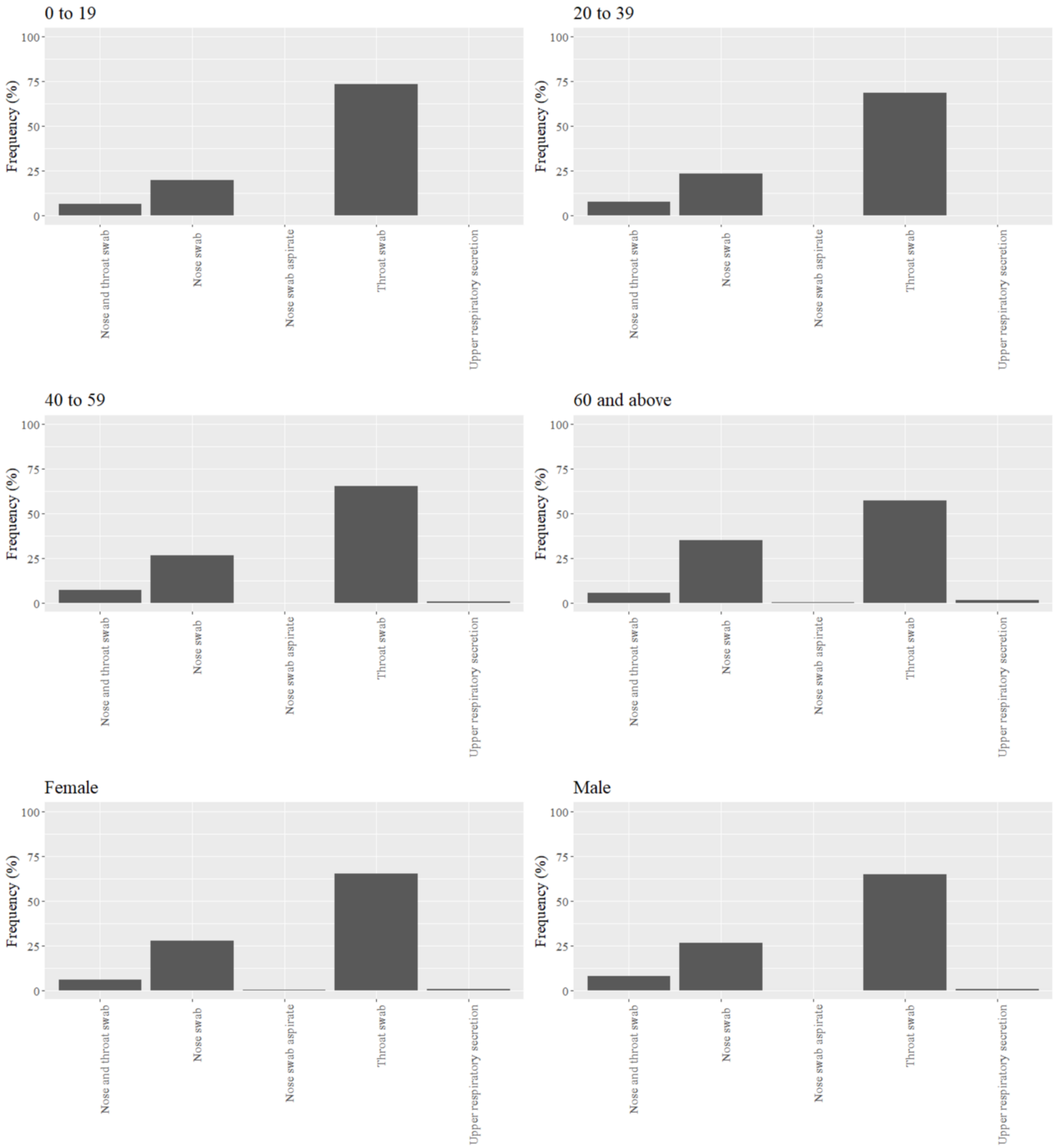


Figure S3. The proportion of genomes isolated using the different upper respiratory tract sample types by age groups and gender in the global population.

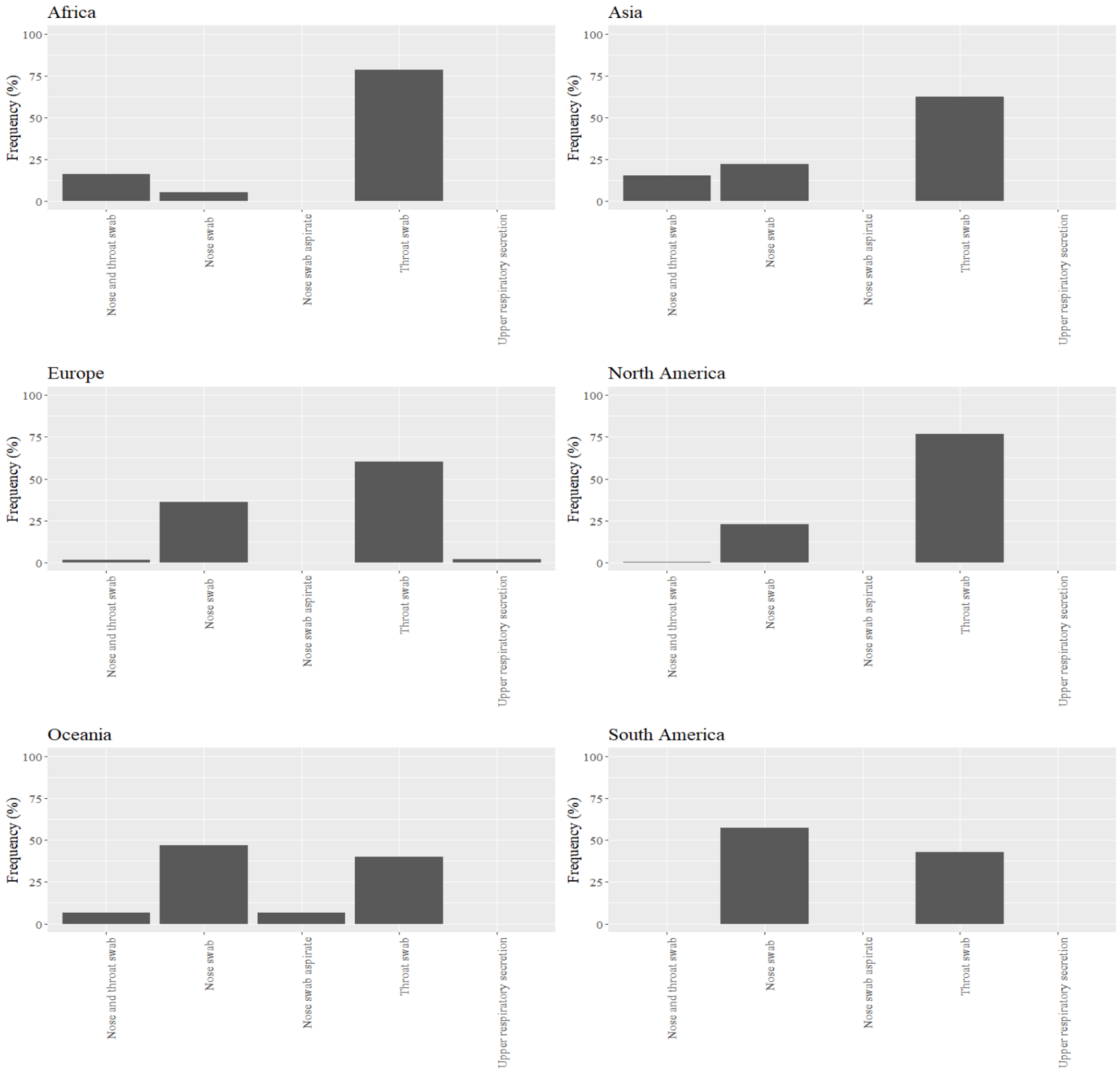


Figure S4. The proportion of genomes isolated using the different upper respiratory tract sample types by continent.

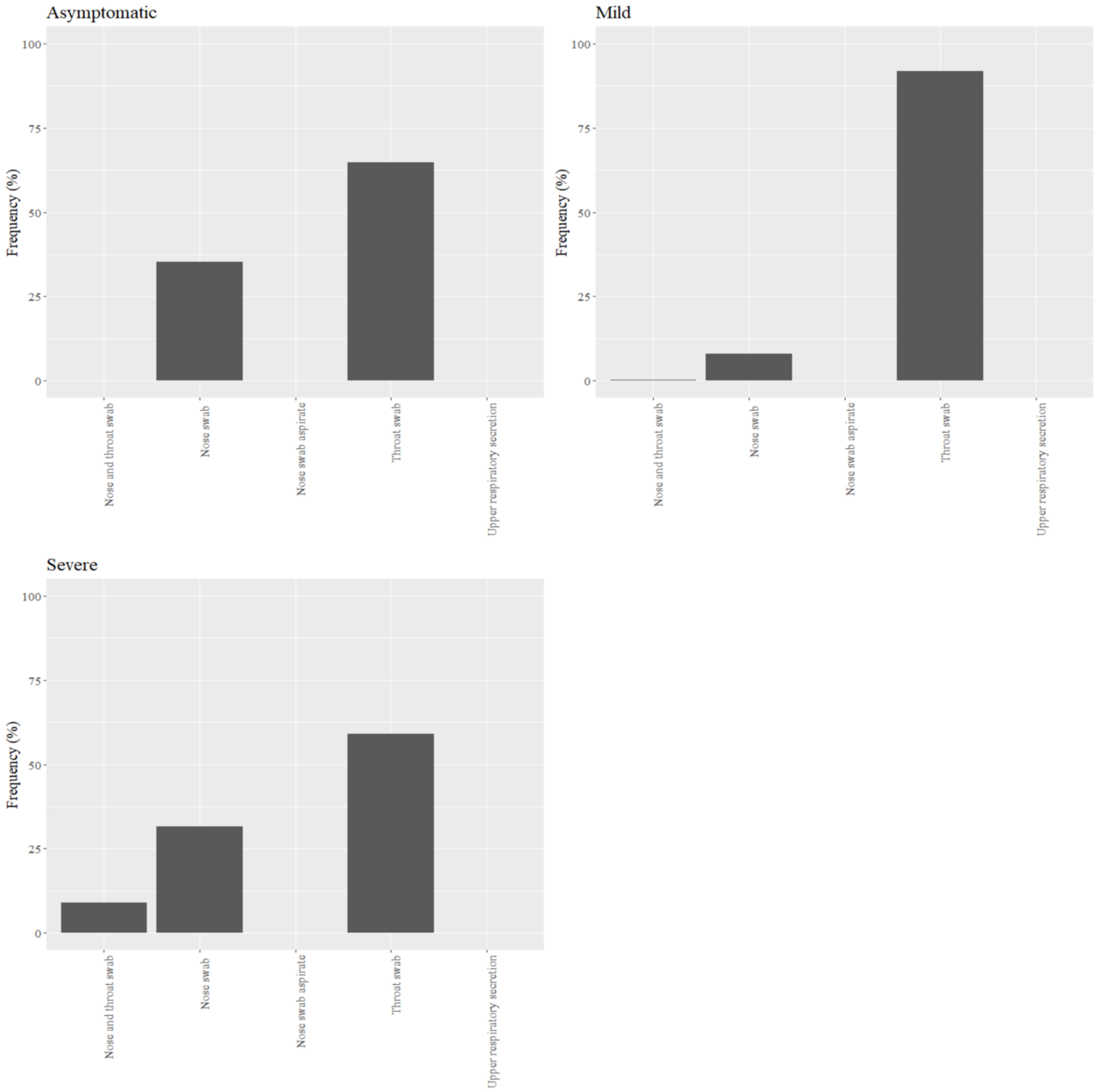


Figure S5. The proportion of genomes isolated using the different upper respiratory tract sample types by COVID-19 disease phenotype in the global population.

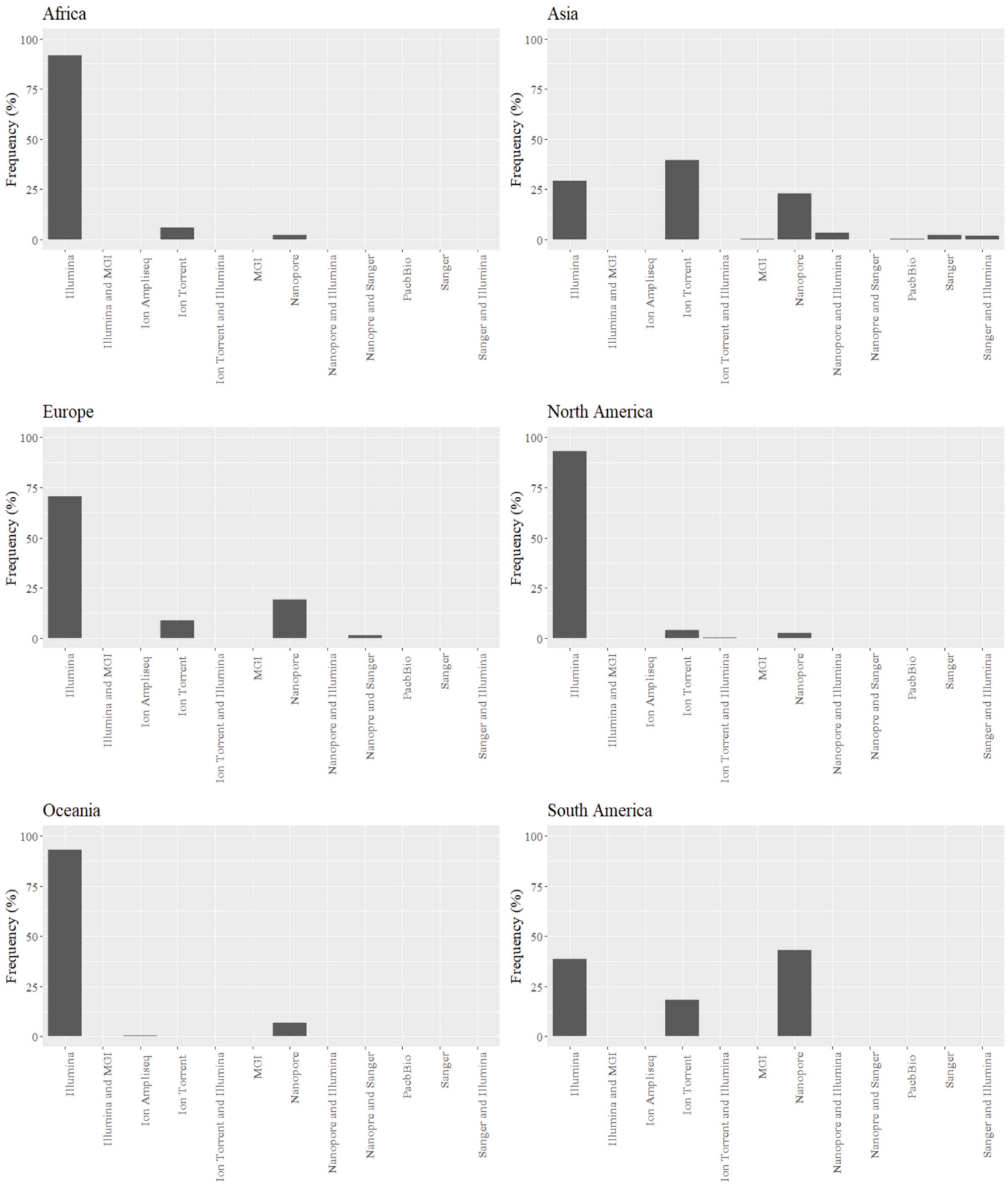


Figure S6. The proportion of genomes sequenced using the different sequencing technologies or platforms is shown for each continent.

Figure S

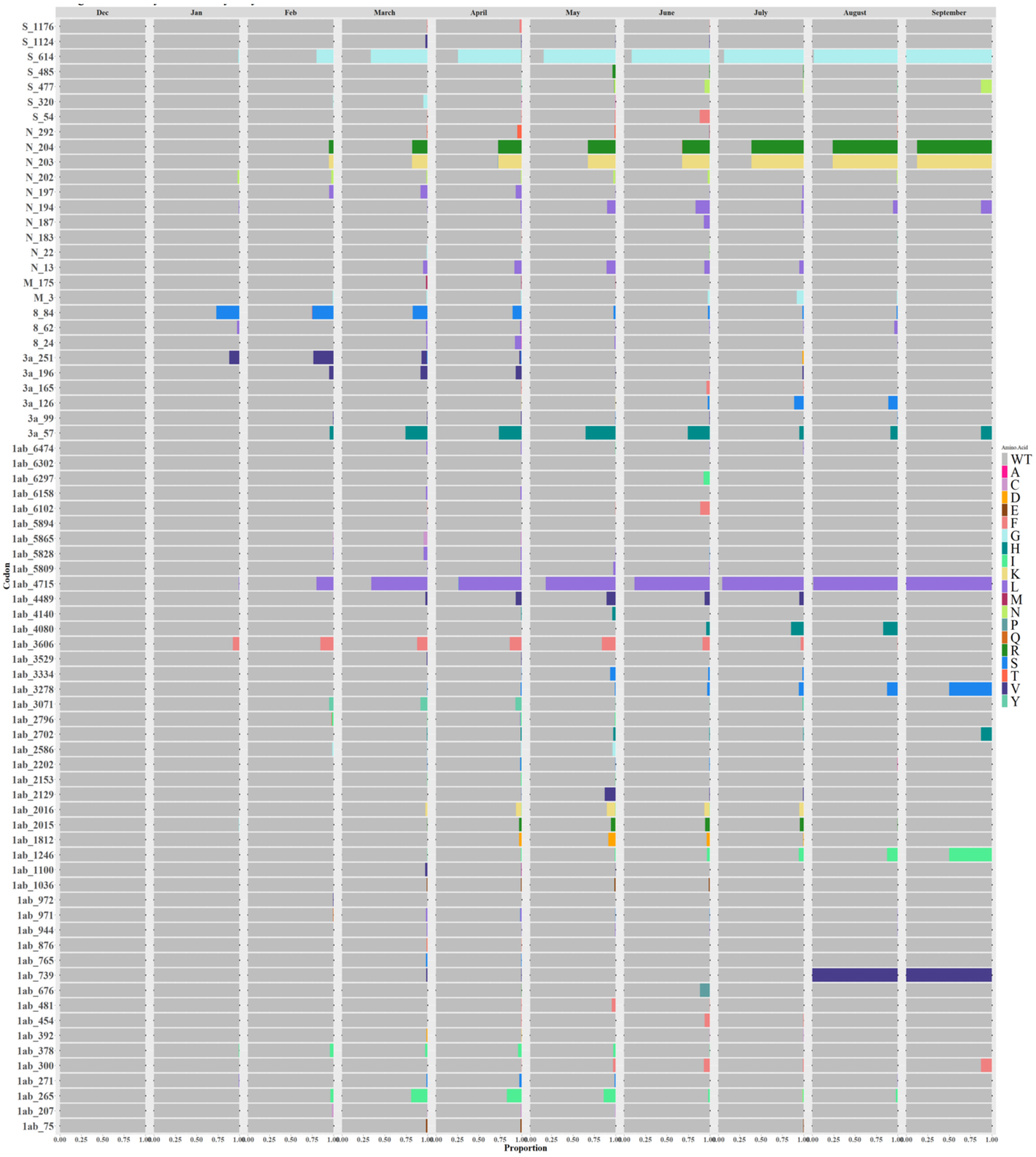


7. Longitudinal prevalence of alleles detected at the 74 loci in the global SARS-CoV-2 population.

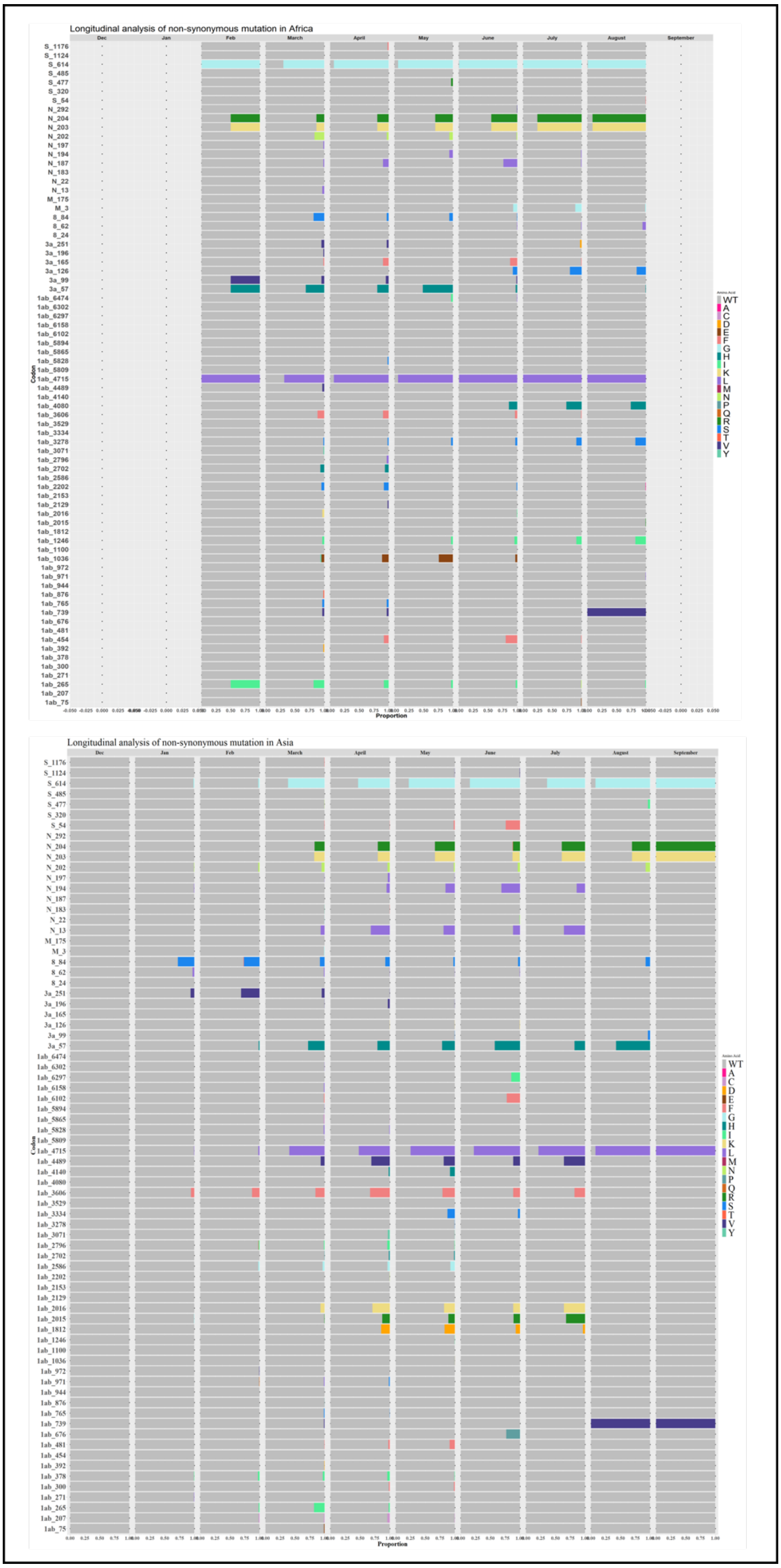


Figure S8. Longitudinal prevalence of alleles detected at the 74 loci in Africa and Asia SARS-CoV-2 populations.

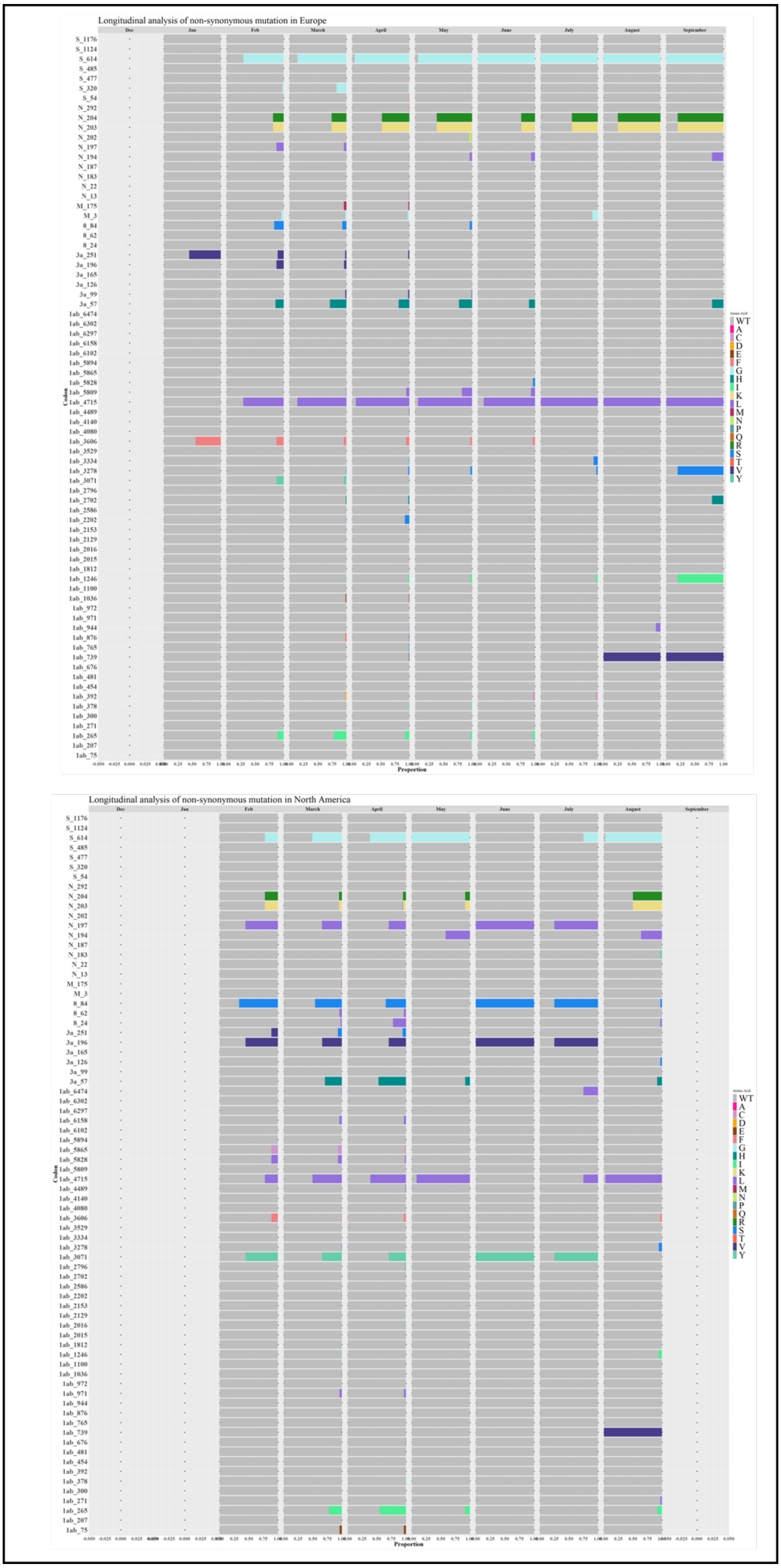


Figure S9. Longitudinal prevalence of alleles detected at the 74 loci in Europe and North America SARS-CoV-2 populations.

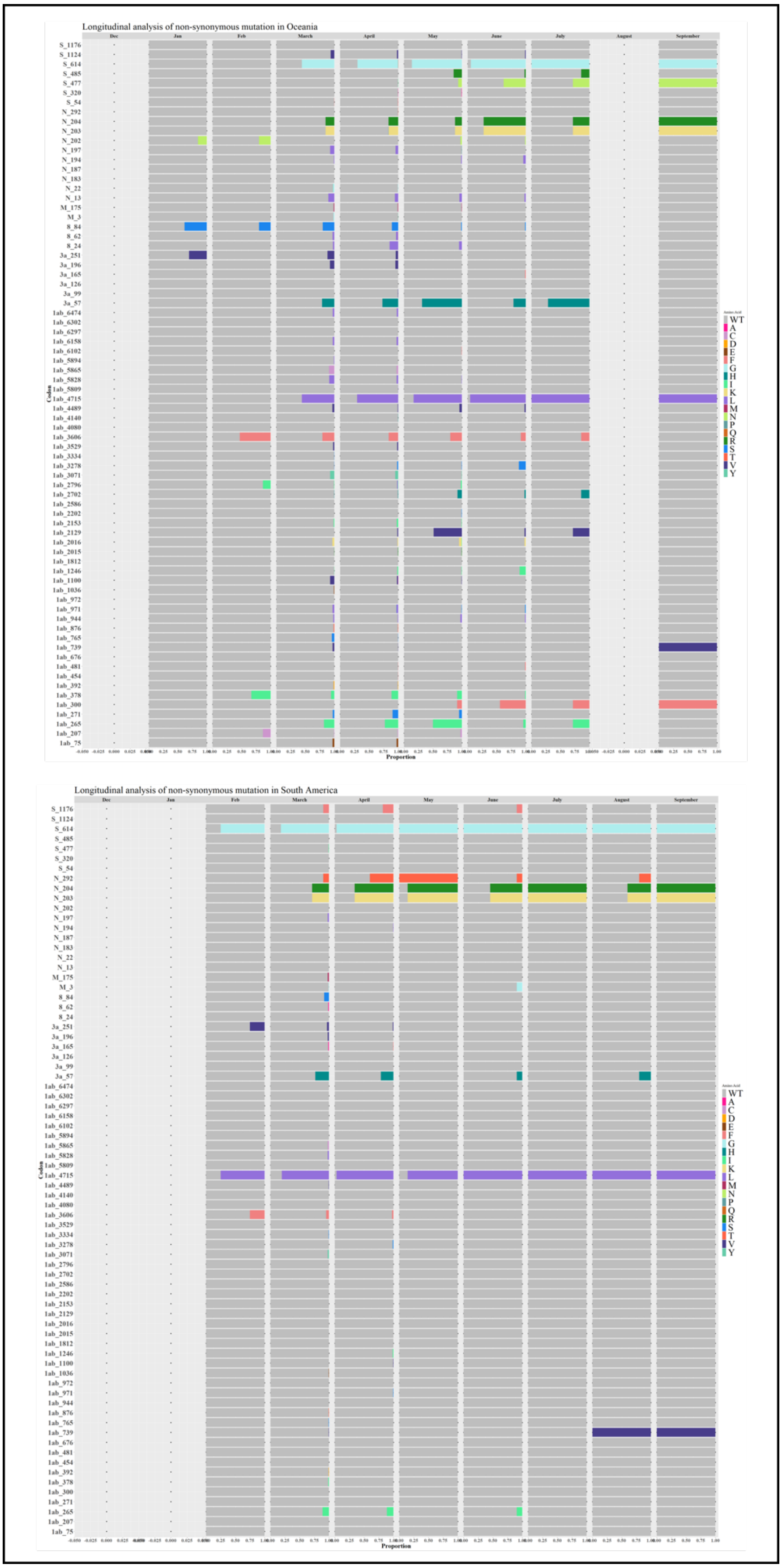


Figure S10. Longitudinal prevalence of alleles detected at the 74 loci in Oceania and South America SARS-CoV-2 populations.

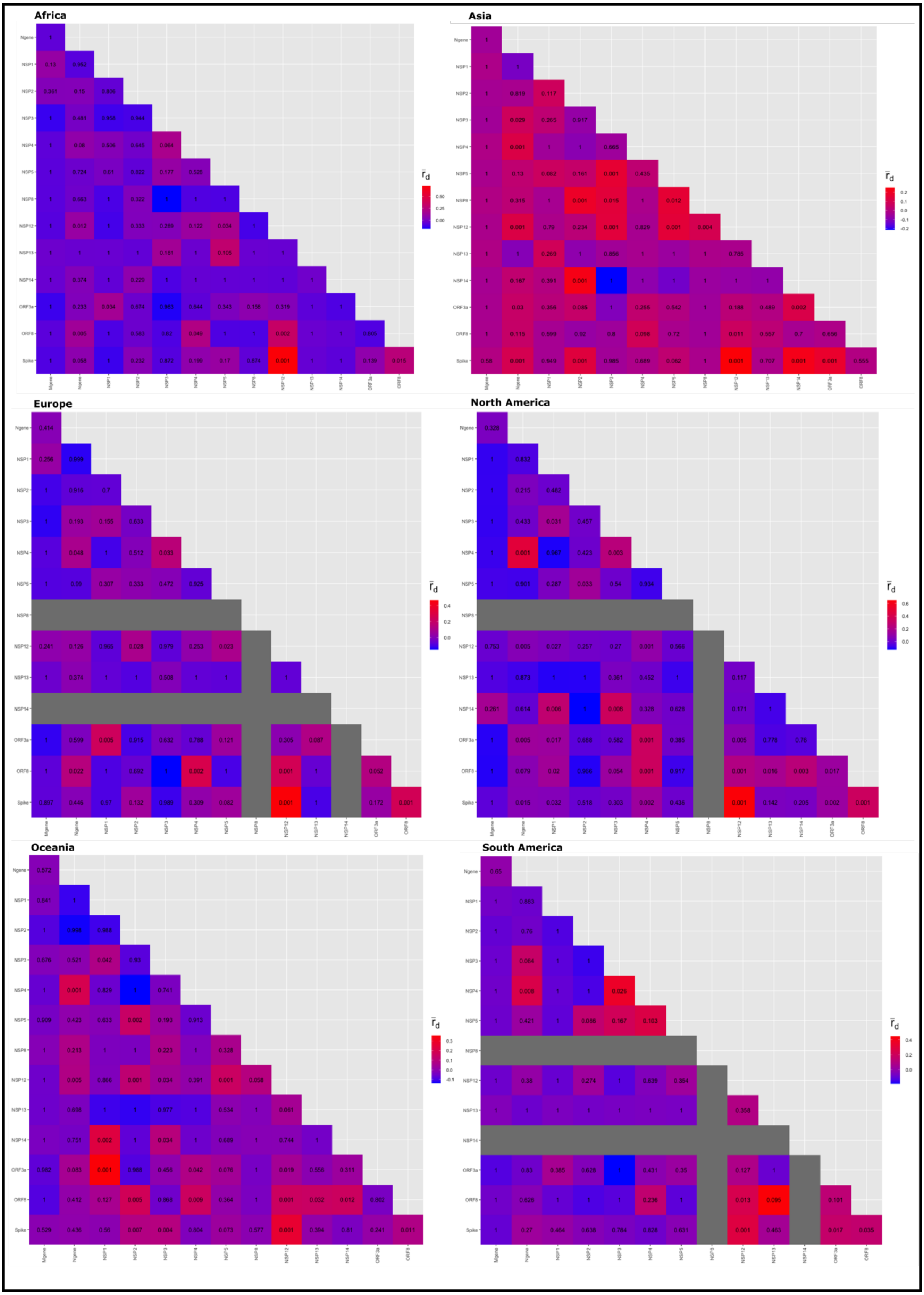


Figure S11. Pairwise LD estimates among genes in the genomes sampled per continent. The $\bar{r}d$ score ranges from 0 (no LD) to 1 (complete LD). The values in the heatmap indicate the P-value associated with the pairwise $\bar{r}d$ estimates. The NSP12 and S LD signal ($\bar{r}d \geq 0.3$, P-value $< 0.001$) was prominent in all continents.

**Africa**

| | North Africa | Central Africa | West Africa |
|---|---|---|---|
| Central Africa | 0.217 | | |
| West Africa | 0.151 | 0.227 | |
| South Africa | 0.327 | 0.374 | 0.216 |

| Genetic Diff. | Gst |
|---|---|
| Little | ≤ 0.09 |
| Moderate | 0.1 - 0.19 |
| Great | ≥ 0.2 |

**Asia**

| | China | Central Asia | East Asia | South Asia | Southeast Asia |
|---|---|---|---|---|---|
| Central Asia | 0.396 | | | | |
| East Asia | 0.135 | 0.392 | | | |
| South Asia | 0.364 | 0.503 | 0.197 | | |
| Southeast Asia | 0.241 | 0.400 | 0.151 | 0.152 | |
| Western Asia | 0.430 | 0.543 | 0.227 | 0.111 | 0.283 |

**Europe**

| | Eastern Europe | Northern Europe | Southern Europe |
|---|---|---|---|
| Northern Europe | 0.248 | | |
| Southern Europe | 0.076 | 0.069 | |
| Western Europe | 0.181 | 0.174 | 0.107 |

Figure S12. Within continent pairwise genetic differentiation (Nei's Gst) among regional blocks in Africa, Asia and Europe.
